# Stewarding scarce response capacity: an inductive qualitative interview study of emergency medical dispatchers prioritising ambulance resources

**DOI:** 10.64898/2026.02.14.26346167

**Authors:** Peter Hill, Daniel Jonsson, Jakob Lederman, Peter Bolin, Veronica Vicente

## Abstract

**Objective:** To explore emergency medical dispatchers (EMD) experiences of prioritising patients and stewarding ambulance resources when system capacity is constrained.

**Design:** Qualitative interview study using inductive qualitative content analysis.

**Setting:** Emergency medical communication centres (EMCCs) in Sweden operated by the national emergency call provider, responsible for receiving 112 calls and dispatching ambulances.

**Participants:** Thirteen purposively sampled EMDs with at least one year of professional experience.

**Data analysis:** Interviews were analysed inductively using qualitative content analysis (Elo and Kyngäs) through open coding, grouping into subcategories and abstraction into generic categories and one main category.

**Results:** Dispatchers described prioritisation under scarcity as system work that simultaneously addresses individual patient acuity and population-level readiness. One main category captured this work: Stewarding scarce response capacity. Three interrelated generic categories characterised stewardship: (1) prioritising by clinical urgency within geographic and operational constraints; (2) producing availability through anticipation, reassessment and queue governance in a ‘virtual waiting room’; and (3) coordinating response through information infrastructures and interprofessional collaboration. Across categories, dispatchers described redistributing risk across patients and time while attempting to avoid both under-response to urgent need and over-allocation that would leave areas without coverage.

**Conclusions:** Dispatch under scarcity is best understood as active stewardship of a safety‐critical dispatch queue. Strengthening patient safety therefore requires organisational support for reassessment and escalation during prolonged waits, and governance that makes queue dynamics and geographic coverage trade-offs visible, rather than relying solely on initial triage decisions or aggregate response-time targets.

**Strengths and limitations of this study:**

- Strengths and limitations of this study
- An inductive qualitative content analysis allowed categories to emerge from dispatchers’ own descriptions, rather than imposing predefined theoretical frameworks.
- Inclusion of emergency medical dispatchers with varied ages, professional experience and EMCC locations enhanced the richness of the data and potential transferability.
- Analyst triangulation, an explicit abstraction pathway and data-to-category quotations strengthened analytic transparency and trustworthiness.
- Interviews were conducted via video, which may have limited access to non-verbal cues compared with in-person interviews.
- The study was conducted within a single national dispatch system, and participation was voluntary, which may limit transferability and introduce self-selection of more experienced or engaged dispatchers.

## Introduction

Emergency medical communication centres (EMCCs) constitute the first formal link in the prehospital emergency care chain. They receive emergency calls, establish the nature and urgency of incidents, and coordinate appropriate responses. In Sweden, calls to the national emergency number (112) are handled by the national provider, which connects callers to emergency medical dispatch. Through early identification of time-critical conditions and timely allocation of resources, EMCCs influence access to emergency care and patient outcomes^1^.

Emergency medical dispatchers (EMDs) verify incident location, elicit salient clinical information, and apply structured dispatch protocols to assign a priority level and response type^2 3^. Their work encompasses call-taking, clinical assessment, and the allocation and dispatch of ambulances. This study, focus specifically on the allocation and dispatch component. Priority levels constitute a shared operational language for urgency (e.g., immediate responses with lights and sirens, urgent responses without lights and sirens, and lower-priority responses that may be safely deferred). However, dispatch is not solely a matter of clinical categorisation. Response decisions are continuously shaped by the real-time availability and geographic distribution of ambulances and other resources, as well as the need to preserve system coverage for sudden high-acuity events^4 5^.

Resource shortages have become a recurrent operational condition in many emergency medical services (EMS) systems, driven by rising demand, prolonged ambulance turnaround times and workforce constraints. When demand exceeds capacity, dispatchers must allocate resources across multiple patients who compete for attention and response. Delays can expose patients to harm through deterioration while waiting, whereas dispatching distant units or over-committing local resources can increase vulnerability elsewhere by leaving geographic areas without immediate response capacity^6^.

Existing research has largely examined scarcity through quantitative indicators such as response times, unit hour utilisation and models for optimal ambulance placement. These approaches can describe system behaviour but provide limited insight into the practical and ethical reasoning required when dispatchers must decide who can safely wait, which risks are acceptable, and how to maintain readiness across a region. Qualitative research has shown that dispatcher decision-making is influenced by call complexity, stress, organisational support and the quality of information elicited during emergency calls^7 8^. Yet much of this research focuses on call assessment or protocol use, rather than on the broader work of coordinating allocation and maintaining system readiness during sustained scarcity^4 5 9 10^.

An inductive qualitative approach can clarify how dispatchers themselves describe and make sense of prioritisation under scarcity, including how they integrate protocol categories with contextual judgement, ethical responsibility and experiential knowledge. Such knowledge may inform training, organisational support and decision-support tool development that aligns with the realities of dispatch work^11^, including the governance of queued cases and the coordination of multiple actors.

### Aim

To explore emergency medical dispatchers’ experiences of ambulance resource shortages and patient prioritisation using an inductive qualitative approach.

## Methods

### Study design and reporting

We conducted a qualitative descriptive interview study. Semi-structured interviews were used for data collection, and inductive qualitative content analysis was used to analyse interview transcripts. This manuscript follows the qualitative content analysis process described by Elo and Kyngäs^12^ (preparation, organising and reporting phases) and is reported in accordance with the Consolidated criteria for Reporting Qualitative research (COREQ)^13^.

### Setting

The study was conducted in Swedish EMCCs operated by SOS Alarm AB, the national emergency call provider responsible for handling 112 calls and coordinating emergency services. EMCCs operate 24/7 and coordinate ambulance dispatch across diverse geographic areas. Ambulance services are delivered by both public and private EMS providers. Dispatchers used a national criteria-based dispatch protocol to assign priority levels and response types, and they coordinated with registered nurses/medical advisors and operational staff who supported resource coordination.

Work is organised to balance call handling and dispatching across centres; dispatchers may therefore manage incidents outside their usual geographic area while maintaining awareness of regional resource availability. In addition to EMDs, EMCC teams may include operational roles that support system-level coordination of ambulances and reassessment of queued cases. In this manuscript, ‘internal officer’ (IB) denotes operational staff involved in ambulance resource coordination.

### Participants, sampling and recruitment

Strategic purposive sampling^12 14^ was used to recruit EMDs with experience of prioritisation during periods of constrained ambulance availability. Inclusion criteria were (1) current employment as an ambulance dispatcher in a Swedish EMCC and (2) at least one year of dispatch experience. We sought variation in sex, age, experience and broad geographic region (northern, central and southern Sweden, including metropolitan Stockholm) to support transferability. No patients or members of the public were involved.

With organisational permission, study invitations describing the purpose, procedures and voluntary nature of participation were distributed internally. Interested dispatchers contacted the research team. Participation or non-participation had no employment consequences, and managers were not informed which individuals participated.

Thirteen EMDs participated (8 women, 5 men; aged 33-62 years). Sample size was guided by information power: the study aim was specific, participants were highly relevant, interviews were in-depth and analysis involved iterative team discussion. Recruitment ceased when no substantively new content emerged that changed the developing category structure. All participants fulfilled the interview.

### Data Collection

Data were collected through in-depth semi-structured video interviews conducted via Microsoft Teams in May 2025. Interviews were conducted by the first and last author (PH (male), VV (female)). Both interviewers have professional experience in EMCC/EMS contexts but had no prior relationship with the participants. Only the interviewer and the participant were present. Interviews lasted a mean of 48 minutes (range 35-62 minutes). Repeat interviews were not conducted.

The interview guide was developed by the research team based on the study aim, relevant literature on emergency medical dispatch and operational experience. The guide was used flexibly and allowed participants to lead the narrative. Interviews began with: “Could you tell me about your experiences working with ambulance dispatching during periods of resource shortages?” Probes explored (a) how priorities were determined when multiple patients competed for resources, (b) how queued cases were managed over time, (c) perceived consequences of delay, and (d) coordination within the EMCC and with external agencies. The full guide is provided in Online supplemental file 1.

All interviews were audio-recorded and transcribed verbatim in Swedish. Transcripts were de-identified and assigned participant numbers. Transcripts were not returned to participants for comment or correction, and no formal participant feedback on preliminary findings was sought. Field notes were written after each interview to capture contextual impressions, emerging topics and reflexive considerations (eg, how interviewer assumptions may have shaped probing). Coding and category development were organised in structured spreadsheets (Microsoft Excel) and text documents.

### Language and translation

Analysis was conducted using Swedish transcripts. Quotations were translated into English by a Swedish English bilingual team member and reviewed by a second bilingual author familiar with EMS terminology. Discrepancies were resolved through discussion with reference to the Swedish original to preserve meaning and nuance.

### Data Analysis

An inductive qualitative content analysis was conducted following Elo and Kyngäs^12^. In line with their description of the preparation phase, PH and VV read each transcript repeatedly to gain a sense of the whole and to understand the context of dispatchers’ experiences. During this familiarisation, we noted preliminary impressions and used field notes to support reflexive awareness of assumptions.

In the organising phase, PH and VV performed open coding line-by-line. Codes were written close to participants’ wording and without a predefined coding frame. We compared codes for similarities and differences, grouped them into subcategories at a predominantly manifest (text-close) level, and then abstracted subcategories into generic categories by capturing shared patterns across participants. Finally, one main category was formulated to express the overarching meaning that integrated the generic categories. Throughout, we moved iteratively between transcripts, codes and evolving categories to refine boundaries and ensure that categories remained grounded in the data.

To distinguish what we did from what the method literature describes, we report our analytic steps (coding, grouping, abstraction, team discussions and audit trail documentation) and cite Elo and Kyngäs^12^ as the methodological source for this analytic approach. Coding sheets and dated versions of category definitions were maintained as part of an audit trail to support dependability and confirmability.

### Trustworthiness and reflexivity

Credibility was supported through purposive sampling of dispatchers with direct experience of scarcity, open-ended interviewing and the use of quotations to demonstrate linkage between data and categories^12 14^. Analyst triangulation was achieved through iterative analytic meetings, review of coding sheets and category definitions by multiple authors, and discussion of alternative interpretations until consensus was reached^14^.

Dependability was strengthened by following a structured content analysis process and documenting analytic decisions over time. Both interviewers’ professional experience in EMCC/EMS contexts supported rapport and context-sensitive probing but also posed a risk of implicit assumptions. To manage this, we used reflexive memoing after interviews and during analysis, explicitly discussed preunderstandings in the author team, and invited scrutiny from co-authors with complementary perspectives to challenge interpretations and category boundaries. Confirmability was supported by systematically linking interpretive claims to data extracts and by searching for deviant cases that challenged the emerging structure^12^. Transferability is supported by a description of the setting and sampling strategy; to protect participant anonymity, detailed site-level characteristics are reported at the level of broad regions rather than specific centres.

### Patient and public involvement

Patients and members of the public were not involved in the design, conduct, reporting or dissemination plans of this research.

## Results

The inductive analysis yielded one main category and three generic categories comprising nine subcategories (Figure 1). To enhance transparency, we describe how the main category integrates and is supported by the generic categories, and we use quotations selectively to illustrate key patterns.

**Figure 1.**
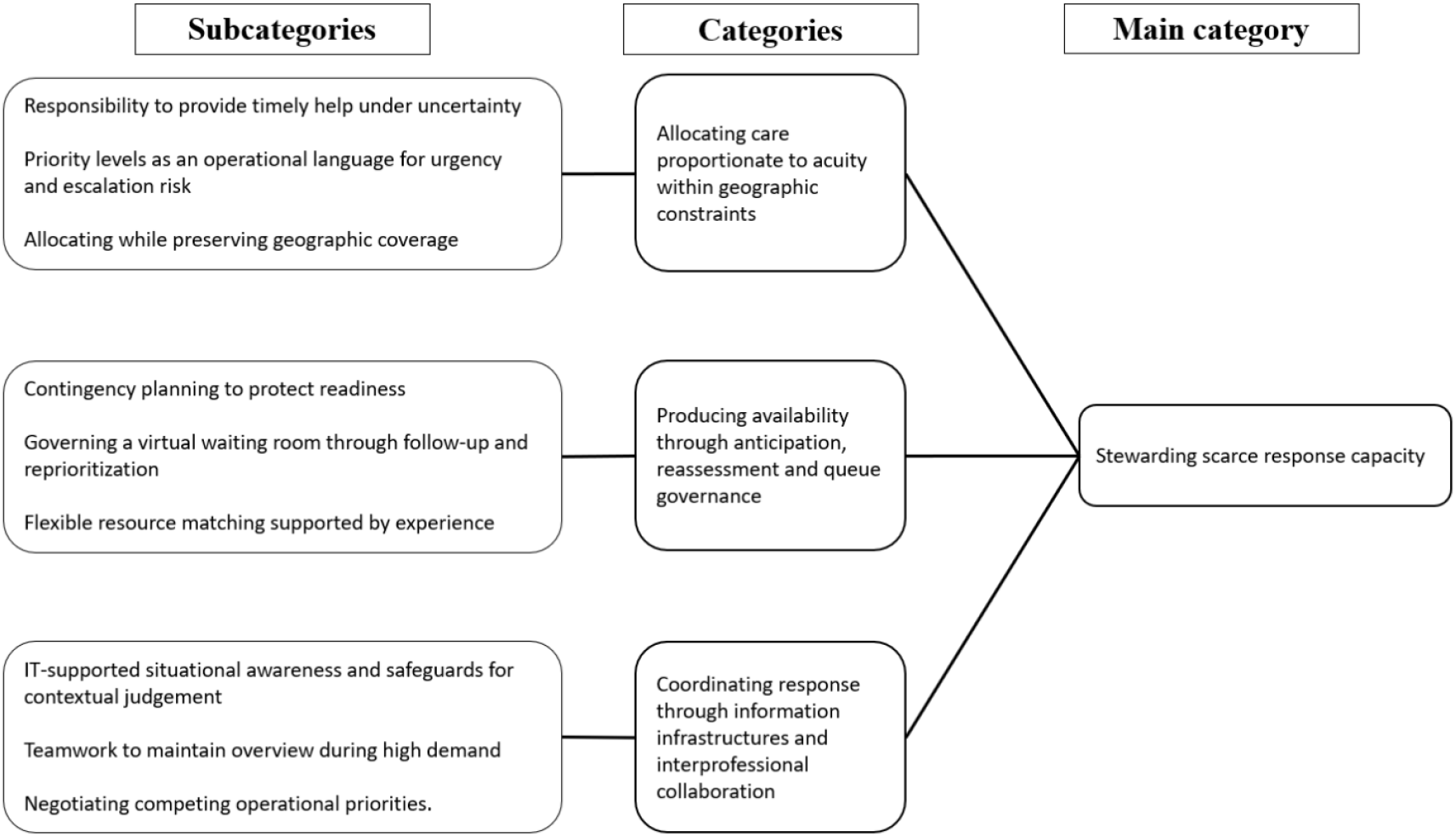
Abstraction pathway from codes → subcategories → generic categories → main category (Stewarding scarce response capacity).

### Main category: Stewarding scarce response capacity

Stewarding scarce response capacity captured dispatchers’ descriptions of sustaining safe service delivery when ambulance availability fluctuated and, at times, was severely constrained. Dispatchers framed their work as stewardship because they were responsible not only for assigning ambulances to individual patients, but also for maintaining readiness for unknown future calls. Stewardship involved redistributing risk: delaying some patients in order to preserve capacity for potentially life-threatening events, and mobilising or repositioning resources to maintain minimum geographic coverage. The generic categories below describe the key forms of work through which this stewardship was enacted.

### Generic category 1: Allocating care proportionate to acuity within geographic constraints

Dispatchers described prioritisation as clinically informed and ethically consequential allocation work carried out in a multi-patient environment. Judgements of urgency were made alongside operational constraints, including distance, travel time, and the system’s need to preserve coverage for sudden high-acuity incidents. Under scarcity, prioritisation was described as selecting not only who should be dispatched first, but also which risks could be accepted at a given moment.

### Responsibility to provide timely help under uncertainty

Dispatchers described a duty to ensure that help was proportionate to need, including anticipating deterioration while patients waited. Ethical reasoning was embedded in routine allocation decisions and intertwined with contextual barriers such as distance, accessibility and uncertainty about the patient’s condition. Participants also described that, during scarcity, striving for an ideal response to every patient could undermine overall safety by exhausting scarce capacity; therefore, they sometimes accepted that lower priorities would exceed nominal time targets in order to protect readiness for time-critical events.

> *“I have the care responsibility. And then I have to make sure the person gets help quickly— how quickly depends on the condition, of course. And where they are… are they out in the forest? Of course that complicates things*.*”*

### Priority levels as an operational language for urgency and escalation risk

Priority levels were described as a shared language that enabled coordination, but dispatchers emphasised that the distribution of priorities across the queue mattered. Cases classified as urgent but not highest priority were described as particularly challenging when multiple such cases accumulated, because they required timely response while ambulances were tied up on simultaneous highest-priority assignments. In these situations, dispatchers described weighing nominal targets against contextual indicators of escalation risk (eg, worsening symptoms, safety concerns or prolonged waiting) and the consequences of allocating a unit from far away.

> *“A 2A is, in our world, a priority 1 without blue lights… it’s almost more of a stress factor when several are waiting*.*”*

### Allocating while preserving geographic coverage

Maintaining minimum coverage across a region was described as a recurrent constraint that shaped allocation. Dispatchers weighed whether to dispatch a distant ambulance immediately or to wait for a nearer unit expected to become available soon, recognising that either choice redistributed risk across patients and time. Some described using planned assignments to move ambulances toward areas with poor coverage while keeping them interruptible for escalation. Geographic reasoning was therefore inseparable from clinical prioritisation in situations where the next high-acuity call could arrive at any moment.

> *“You have to think what is actually fastest for the patient: assign an ambulance with a 30-minute drive… or wait for a unit that will soon be available nearby*.*”*

### Generic category 2: Producing availability through anticipation, reassessment and queue governance

Dispatchers described actively producing readiness rather than merely responding to incoming calls. This work included contingency planning, continual regulation of a ‘virtual waiting room’ and experience-based strategies to retain control when demand exceeded available ambulances. Readiness was described as a fragile system property that could improve or collapse within minutes, requiring proactive planning and continual reprioritisation.

### Contingency planning to protect readiness

Preparedness was described as continuous replanning and repositioning to protect response capacity as conditions changed. Dispatchers described monitoring where ambulances were likely to become available, anticipating peaks in demand and preparing alternative options (eg, engaging other rescue resources) when ambulances were unavailable. This planning was experienced as essential for meeting time-critical needs despite volatility.

> *“You can’t just have a plan A. You need a plan B, C and so on*.*”*

### Governing a virtual waiting room through follow-up and reprioritisation

Participants described a structured queue for cases that could not be dispatched immediately, often referred to as a virtual waiting room. Governance of this queue involved monitoring, reassessment and reprioritisation to manage clinical risk over time. Dispatchers described relying on reassessment processes (including support from nurses/medical advisors) to detect changes in symptoms and trigger escalation when warranted. They also described how queue length could grow rapidly during simultaneous high-priority events or when ambulances were temporarily unavailable, shifting the work from first-come-first-served dispatch to continuous risk management across waiting patients.

> *“The waiting room is all healthcare calls that are not a priority 1… you check: has anything changed? Do we need to change priority?”*

### Flexible resource matching supported by experience

Dispatchers described allocation as matching resource type, competence and response mode to the situation rather than assigning a priority alone. This included deciding when specialised resources were appropriate and when they were too distant to be clinically reasonable. Experience was portrayed as stabilising: it supported pattern recognition, anticipation of bottlenecks, and the use of alternative resources when ambulances were unavailable. Experience was also described as increasing tolerance for uncertainty and reducing reactive over-allocation that could destabilise coverage.

> *“It’s experience… you have ambulances, but you also have resources like the fire service… and helicopters*.*”*

### Generic category 3: Coordinating response through information infrastructures and interprofessional collaboration

Dispatchers’ stewardship relied on information infrastructures that made resource status visible and on collaboration across roles and agencies to manage concurrency. Participants described coordination as both technical and social: it depended on functional systems that supported situational awareness and on teamwork that distributed tasks and aligned decisions under time pressure.

### IT-supported situational awareness and safeguards for contextual judgement

Real-time status monitoring and time tracking supported anticipatory planning by helping dispatchers predict when units would become available and prepare allocations accordingly. Conversely, unexpected time extensions disrupted plans and could cascade across the queue. Dispatchers valued automation and rapid dispatch functions for speed, but emphasised that some call types required contextual listening and should be routed for human judgement to prevent unsafe automatic dispatch.

> *“There are some not included—psychiatry, suicide, assault—where you may need to listen. It’s not good that it goes out automatically directly*.*”*

### Teamwork to maintain overview during high demand

Collaboration within the EMCC helped distribute tasks and maintain shared situational awareness when workload was high. Participants described how communication loops and shared monitoring supported rapid updates and enabled dispatchers to focus on the most time-critical tasks while colleagues helped maintain oversight of queued cases.

> *“We collaborate a lot… the ‘loop’ helps… when it’s busy, because you don’t have time to keep up*.*”*

### Negotiating competing operational priorities

Dispatchers sometimes experienced tension between patient-centred urgency and operational considerations such as breaks, staffing or workload distribution. While acknowledging the need for staff endurance and safe working conditions, participants described frustration when operational routines were perceived to delay response for patients who had already waited a long time. Such situations required negotiation and, at times, escalation to ensure that allocation decisions remained aligned with patient safety while sustaining the system over prolonged demand.

> *“Sometimes you feel frustrated that someone has to wait a long time so the crew can sit and eat their lunch boxes in peace*.*”*

## Discussion

### Principal findings

This study explored emergency medical dispatchers’ experiences of prioritising patients when ambulance capacity is constrained. Dispatchers described prioritisation under scarcity as stewardship of response capacity: a form of system work that integrates acuity assessment with geographic coverage, queue governance, anticipation and coordination across roles. Rather than portraying prioritisation as a linear application of protocol categories, participants described continuously redistributing risk across patients and time while trying to avoid both under-response to urgent need and over-allocation that would compromise readiness for future time-critical events. Interpreted alongside prior work, this supports the view that structured dispatch tools provide a foundation, but that experiential and contextual judgement remain central when cases are complex or scarcity intensifies^11^.

### Stewardship as system work and ethical-operational responsibility

The main category, stewardship, captures the dual accountability dispatchers described: responsibility for individual patients in the queue and responsibility for maintaining a functioning emergency response system. This dual accountability rendered prioritisation inherently ethical-operational. Dispatchers described how decisions about who could wait were shaped by judgements about clinical urgency, uncertainty and potential deterioration, but also by the recognition that using the ‘last’ available ambulance could increase risk for unknown future calls. In this sense, dispatch prioritisation under scarcity can be understood as system-level clinical reasoning: it integrates structured triage tools with contextual judgement about geography, time, and downstream consequences of delay.

Importantly, stewardship was not described as a general attitude but as practical work enacted through specific activities: preserving geographic coverage, monitoring and reprioritising waiting cases, mobilising alternative resources and coordinating with colleagues to maintain overview. Making these activities visible may help move organisational discussions beyond individual ‘good’ or ‘bad’ dispatch decisions to a recognition of the infrastructure and support required to enact safe stewardship.

### Queue governance and reassessment as a safety mechanism

A central contribution of the findings is the depiction of a ‘virtual waiting room’ as an active safety practice rather than a passive backlog. Quantitative and modelling work has examined EMS demand and performance through measures such as response times, workload and system behaviour^4 5 10^. While such studies improve understanding of time-dependent patterns, they provide limited insight into how scarcity is experienced and managed moment-to-moment in the dispatch setting. In this study dispatchers described reassessment and reprioritisation as essential for detecting change over time, particularly when waiting times extend beyond nominal targets. This suggests that patient safety under scarcity depends not only on the initial priority assignment but also on the capacity to sustain follow-up work across waiting patients. Such capacity requires staffing, clear routines, and decision-support that makes deteriorating risk visible. Without these, queued patients may become ‘invisible’, and escalation may be detected late. This findings complement existing evidence^7 8^.

### Geographic coverage and equity-oriented trade-offs

Dispatchers’ emphasis on preserving geographic coverage illustrates how equity and readiness constraints enter everyday allocation work. Dispatch and EMS performance literature emphasising timeliness and resource allocation as key system concerns^1 4 5^. Sending a distant unit may reduce delay for one patient but increase vulnerability for others by leaving areas uncovered. Conversely, waiting for a nearer unit may preserve coverage but prolong the exposure to risk for the patient waiting. These trade-offs were described as unavoidable under scarcity. Organisational guidance that explicitly addresses such trade-offs could support consistency, reduce moral distress and provide a shared language for evaluating decisions when ‘ideal’ responses are not feasible.

### Information infrastructures, automation and collaboration

Dispatchers described information infrastructures as integral to coordination. This is consistent with qualitative dispatch literature emphasising the role of information quality and organisational context in decision-making^7 8^. Status monitoring and time tracking supported anticipatory planning, while unexpected delays disrupted plans and could cascade across the queue. Participants welcomed automation that increased speed, but emphasised that automation must include safeguards for call types that require contextual listening, such as safeguarding concerns or complex psychiatric emergencies. The findings therefore support a principle of ‘selective automation’: rapid functions should be combined with routings that preserve human judgement when nuance is clinically or ethically consequential.

Collaboration within the EMCC and with external agencies functioned as a resilience mechanism. Teamwork helped maintain overview when workload was high, and collaboration with partner agencies was used to mitigate risk when ambulances were unavailable. However, collaboration also involved negotiating competing operational priorities. Interpreted in relation to existing qualitative dispatch evidence, this underscores that coordinated management under scarcity is not only technical but also relational and organisational^7 8^.

### Implications for practice and policy

First, organisations should treat scarcity management as core dispatch work. Training and local routines should explicitly include contingency planning, geographic reasoning and queue governance, including reassessment routines and escalation thresholds for waiting patients. Second, ethically difficult trade-offs should be supported through structured peer consultation, debriefing opportunities and feedback mechanisms that focus on learning rather than individual blame. Third, resilience planning should include redundancy and rehearsed fallback procedures for degraded IT conditions, given the centrality of information infrastructures for coordination. Fourth, decision-support and automation should be designed to increase speed without bypassing contextual judgement for high-risk call types. Finally, mentorship and structured case review may help newer dispatchers acquire the experience-based judgement described as stabilising under scarcity.

### Strengths and limitations

A strength of this study is the inductive analytic approach, which allowed categories to emerge from dispatchers’ accounts without imposing a predefined framework^14^. The sample represented multiple regions within the Swedish dispatch system and included variation in age and experience. Limitations include voluntary participation, video-based data collection, and conducting the study within a single national provider^9^. Interviewers’ professional EMCC/EMS experience may have influenced data collection and interpretation; reflexive memoing, documented audit trail work and multi-author scrutiny were used to mitigate this^14^.

## Conclusion

Emergency medical dispatchers described prioritisation under ambulance scarcity as stewardship of response capacity: system work that balances patient acuity, waiting-time risk and geographic readiness through anticipation, reassessment and coordinated collaboration. Supporting dispatchers’ stewardship requires organisational structures and decision-support that make trade-offs visible, protect contextual judgement and sustain follow-up of patients who must wait.

## Supporting information

COREQ checklist

## Data Availability

Due to confidentiality and privacy considerations, de-identified interview transcripts are not publicly available.

## Acknowledgements

The authors thank the emergency medical dispatchers and EMCC management at SOS Alarm AB for facilitating recruitment and sharing their experiences.

## Notes

### Competing Interest Statement

The authors have declared no competing interest.

### Funding Statement

This study did not receive any funding

### Author Declarations

Swedish Ethical Review Authority (Dnr 2022-03701-01; 13 September 2022)

