## Supplementary material for "Stewarding scarce response capacity: an inductive qualitative interview study of emergency medical dispatchers prioritising ambulance resources": COREQ checklist

**COREQ checklist (Consolidated criteria for Reporting Qualitative research): 32-item checklist**

**Manuscript title:** Allocating the right ambulance to the right patient under resource shortages: an inductive qualitative interview study of emergency medical dispatchers

Page references correspond to the PDF-rendered manuscript version. Where an item is not addressed in the manuscript, it is marked as "Not reported" (in accordance with COREQ guidance).

| Item No. | Item | Guide questions/description | Reported on page # |
|---|---|---|---|
| **Domain 1: Research team and reflexivity** | | | |
| ***Personal characteristics*** | | | |
| 1 | Interviewer/facilitator | Which author(s) conducted the interview? | p.7 |
| 2 | Credentials | What were the researcher's credentials (e.g., PhD, MD)? | p.1 |
| 3 | Occupation | What was the researcher's occupation at the time of the study? | p.1 |
| 4 | Gender | Was the researcher male or female? | Not reported |
| 5 | Experience and training | What experience or training did the researcher have? | pp.7-8 |
| ***Relationship with participants*** | | | |
| 6 | Relationship established | Was a relationship established prior to study commencement? | p.7 |
| 7 | Participant knowledge of the interviewer | What did participants know about the researcher (e.g., goals, reasons for doing the research)? | p.6 |
| 8 | Interviewer characteristics | What characteristics were reported about the interviewer (e.g., bias, assumptions, reasons/interests in the topic)? | p.8 |
| **Domain 2: Study design** | | | |
| ***Theoretical framework*** | | | |

| | | | |
|---|---|---|---|
| 9 | Methodological orientation and theory | What methodological orientation underpinned the study (e.g., grounded theory, discourse analysis, ethnography, phenomenology, content analysis)? | p.6 |
| ***Participant selection*** | | | |
| 10 | Sampling | How were participants selected (e.g., purposive, convenience, consecutive, snowball)? | p.6 |
| 11 | Method of approach | How were participants approached? | p.6 |
| 12 | Sample size | How many participants were in the study? | p.6 |
| 13 | Non-participation | How many people refused to participate or dropped out? Reasons? | p.6 |
| ***Setting*** | | | |
| 14 | Setting of data collection | Where was data collected? | p.7 (video via Microsoft Teams; physical location not specified) |
| 15 | Presence of non-participants | Was anyone else present besides the participant(s) and researcher? | p.7 |
| 16 | Description of sample | What are the important characteristics of the sample (e.g., demographic data, date)? | pp.6-7 (Table 1) |
| ***Data collection*** | | | |
| 17 | Interview guide | Were questions, prompts, guides provided? Was it pilot tested? | p.7 (guide described; provided as Online supplemental file 1; pilot testing not reported) |
| 18 | Repeat interviews | Were repeat interviews carried out? If yes, how many? | p.7 |

| | | | |
|---|---|---|---|
| 19 | Audio/visual recording | Did the research use audio or visual recording to collect the data? | p.7 |
| 20 | Field notes | Were field notes made during and/or after the interview or focus group? | p.7 |
| 21 | Duration | What was the duration of the interviews or focus group? | p.7 |
| 22 | Data saturation | Was data saturation discussed? | pp.6-7 (information power; stopping criterion described) |
| 23 | Transcripts returned | Were transcripts returned to participants for comment and/or correction? | Not reported |
| **Domain 3: Analysis and findings** | | | |
| ***Data analysis*** | | | |
| 24 | Number of data coders | How many data coders coded the data? | p.8 |
| 25 | Description of the coding tree | Did authors provide a description of the coding tree? | pp.8-9 (abstraction pathway + Figure 1) |
| 26 | Derivation of themes | Were themes identified in advance or derived from the data? | p.8 (derived inductively) |
| 27 | Software | What software, if applicable, was used to manage the data? | Not reported |
| 28 | Participant checking | Did participants provide feedback on the findings? | Not reported |
| ***Reporting*** | | | |
| 29 | Quotations presented | Were participant quotations presented to illustrate the themes/findings? Was each quotation identified? | pp.9-14 |
| 30 | Data and findings consistent | Was there consistency between the data presented and the findings? | pp.9-14 |
| 31 | Clarity of major | Were major themes clearly | pp.9-14 |

| | | | |
|---|---|---|---|
| | themes | presented in the findings? | |
| 32 | Clarity of minor themes | Is there a description of diverse cases or discussion of minor themes? | p.8 (deviant/negative cases sought during analysis; minor themes not separately presented) |
